# Predictors of gestational age at birth and its association with postnatal growth outcomes in preschool-age children

**DOI:** 10.1101/2025.05.01.25326790

**Authors:** Mahama Saaka, Comfort Kuulabio

## Abstract

**Background:** Although the length of gestation has been shown to have an effect on pregnancy outcomes including birth weight, information regarding its effect on postnatal growth of children is scanty. The aim of this study was to assess the potential factors affecting duration of gestation, and how it associates with postnatal growth outcomes of children under two years.

**Methods:** This was an analytical health facility-based, cross-sectional study. A systematic random sampling technique was used, to select mother–child pairs who sought services from child welfare clinic (CWC) in the Tamale Teaching Hospital. The predictors of gestational age were identified using multivariable linear regression and hierarchical multivariable linear regression analysis was used to test the independent contribution of gestational age to length-for-age z-score (LAZ).

**Results:** The prevalence of late term/post-term delivery was 75 (23.5%). Older women were positively associated with longer gestational age as a unit increase in age of the mother increased gestational age by 0.28 standard units, β = 0.28, (95% CI: 0.07 to 0.20). Malarial infection during pregnancy reduced duration of gestation by 0.11 standard units, β = -0.11, (95% CI: -0.79 to -0.02). Greater weekly gestational rate of weight gain reduced gestational age by 0.12 standard units, β = - 0.12, (95% CI: -0.005 to -0.01)

An increase in one week of gestational age was associated with 0.16 increase in LAZ [beta coefficient (β), = 0.16 (95% CI: 0.09 to 0.36)]. However, wasting was 2 times more common among the children born late/post term, AOR= 2.18 (95% CI: 1.25, 3.82, and p = 0.006), compared to children born at term (37-40 weeks).

**Conclusions:** After rigorous adjustment for confounding factors, post-term pregnancy was associated with a higher risk of thinness but protected against stunting in preschool-age children. These findings suggest that post-term pregnancies may have complex and potentially conflicting effects on the long-term growth and development of children.

## Introduction

The prevalence of stunted growth among children under two years is unacceptable high in many countries and about 165 million children younger than 5 years old are reported to be stunted [1]. In the Northern Region of Ghana, many children have an inadequate nutritional status as 29.6 %, 7.8 % and 19.9 % of children under five are stunted, wasted and underweight, respectively [2].

Globally, measures are consistently being sought to address child stunting because of its associated negative health and economic outcomes, including shorter adult height, less schooling, and reduced adult income [3]. For example, the WHO in 2012 adopted a resolution on maternal and child undernutrition, targeting a reduction of stunting by 40% by 2025 [4]. The reduction of all forms of malnutrition including stunted growth by 2030 is one of the targets as contained in the United Nations Sustainable Development Goals (SDGs) [5].

Available evidence suggests that growth faltering in the first 1,000 days of life may adversely influence health and human capital and this can be irreversibly [6, 7]. In low resource settings in particular, stunting is influenced by several factors including low birth weight, short maternal stature, low educational level of mothers, poverty, inappropriate breastfeeding, poor infant and young child feeding practices, infections, and other environmental exposures [1, 8–11].

Late/ post term pregnancy (gestational age ≥ 41 weeks ) [12] is reported to be associated with other adverse perinatal and maternal outcome [13, 14] outcomes. Gestational age appears to be non-modifiable factor and little is however, documented on how long duration of pregnancy relates to postnatal growth outcomes.

There’s an observed increase in women delivering at or beyond 41 weeks of gestation at the Tamale Teaching Hospital in Ghana’s Northern Region, which warrants further investigation and potential interventions to ensure maternal and fetal well-being. There is however, little documented on the long-term effect of post-term gestation on postnatal child growth. Furthermore, little is known about the factors that affect the duration of pregnancy. This study therefore sought to understand the potential factors affecting gestational age at birth and its association with postnatal growth outcomes of preschool-age children. Addressing these gaps is essential for informing public health strategies and improving long-term outcomes for children born post-term.

## Materials and methods

### Study design, population and sampling

An analytical health facility-based cross-sectional study design was used to collect data from mothers with children 6-23 months. The study population comprised mother–child pairs who sought services from child welfare clinic (CWC) in the Tamale Teaching Hospital.

At the health facility, systematic random sampling was applied to extract the required sample. The sampling frame was the list of women contained in the postnatal care (PNC) register. On each PNC day, the mothers who met the inclusion criteria were systematically selected. This was done until desired sample size was achieved.

### Sample size calculation

The minimum sample size was estimated based on a single population proportion formula with the assumption of 95% CI and 0.05 margin of error. The main outcome of interest was stunted growth whose population proportion is estimated to be 29.8 % among children in the study area [2]. Based on these parameters, the minimum sample size was estimated to be 322.

### Data collection tools and measurements

Data were collected in face-to-face interviews using structured and pretested questionnaires, which consisted of sociodemographic characteristics, gestational information, and other related variables. The questionnaire was developed by authors after reviewing relevant literature on the topic. The questionnaire was used to collect data on background information, household economic status, ANC/CWC attendance, mode of delivery, birth weight, gestational age and infant and young child feeding practices among others. Anthropometric measurements were made on the weight and length of children.

### Study variables and measurement

The main exposure variable of this study was gestational age (GA), which is defined in weeks as the duration of pregnancy. This was determined using trans-abdominal ultrasound techniques in the hospital each woman attended for antenatal services. Gestation less than 37 weeks was classified as preterm, term pregnancy (≥37 and <41 weeks) which was set as the comparison group, late pregnancy (> 37 and ≤ 41 weeks) and post-term (prolonged) pregnancy was defined as maternal gestational age ≥42 weeks. Therefore, late/ post term pregnancy was gestational age ≥ 41 weeks [12].

### Assessment of gestational weight gain

Gestational weight gain (GWG) was determined as the difference between the weight of each woman in the third trimester (that is, approximately 36 weeks gestation) and weight in the first trimester (that is, in the first 12 weeks of gestation). The mean maternal GWG per week was calculated by dividing the total weight gain in grams by the gestational ages in weeks, making it independent of the pregnancy duration. The revised American Institute of Medicine (IOM) guidelines were used to categorize weekly maternal GWG into inadequate, normal or excessive weight increase per week : within the first quartile (Q1: <419.4 g/week), between the second and third quartile (Q2–Q3: 419.4–692.3 g/week) and within the fourth quartile (Q4: >692.3 g/week) [15, 16].

### Assessment of maternal anthropometry

The mother’s height was measured using a height measure to the nearest 0.1 cm. Weight was measured without shoes and heavy clothing to the nearest 100 g. Body mass index (BMI) was calculated by dividing the body mass (in kilograms) by the square of the body height (in meters). Maternal BMI in early pregnancy (not more than 12 weeks gestation) was also calculated as a proxy for pre-pregnancy BMI [17, 18]. The maternal BMI in early pregnancy was categorized according to the WHO cut off points: underweight (<18.5 kg/m^2^), normal weight (18.5–25.0 kg/m^2^), overweight (25.0^+^–30.0 kg/m^2^ ) and obesity >30 kg/m^2^

### Postnatal growth assessment of children

The primary outcome of interest was post-natal child growth as measured by length-for-age z-score (LAZ) and weight-for-length z-score (WLZ), and weight-for-age z-score (WAZ) and these were calculated as recommended by the World Health Organization [19]. The continuous z-scores were also categorized as short for their age or stunting (LAZ < –2 SD), acutely malnourished or wasting (WLZ < –2 SD) and underweight (WAZ< –2 SD) from the median of the reference population.

### Data quality management

Several measures were taken to ensure the accuracy and reliability of the data collected. The data collectors were trained for two days on the objectives and methodology of the study, relevance of the study, ethical concern, and techniques of interviews. There was pre-testing of the questionnaire during the training. In all 5% of the study questionnaires were pretested and validated with mother- pair child who were not part of the study. Field supervisors checked for completeness and consistency in the field and during data entry to ensure data quality.

### Data analysis

The Statistical Package for Social Sciences (SPSS version 22) was used in the data analysis. The predictors of gestational age were identified using multivariable linear regression and hierarchical multivariable linear regression analysis was used to test the independent contribution of duration of pregnancy (gestational age) to length-for-age z-score (LAZ). The covariate predictor variables (main effects) were entered in the first step of the regression analysis. The covariates included sociodemographic characteristics (maternal age at delivery, maternal educational level, maternal employment type), mothers’ characteristics (weight and height, BMI at recruitment, gravidity, parity, obstetric and medical history of mothers during pregnancy), as well as child characteristics (birth weight, sex, mode of delivery, malarial infection, feeding practices). In the second step, the main explanatory variable of interest (that is, gestational age) was added. The percentage of variability in the dependent variable that could be accounted for by all the predictors together was measured by R-square. The change in R^2^ is a way to evaluate how much predictive power was added to the model by the addition of another variable. So, in step 2 when the explanatory variable was added to the model, the % of variability accounted for was evaluated. The regression analyses are presented as β coefficients with 95%confidence intervals.

The presence of multi-collinearity was checked using variance inflation factors (VIF), but there was no variables whose VIF was more than 2, an indication of no multicollinearity among predictors.

Before the analysis, children who had extreme values of anthropometric z-score values outside the following intervals are considered implausible according to the WHO criteria [20]: HAZ (-6, +6), WHZ (-5, +5), WAZ (-6, +5), BMIfor-age z score >5 or <−5 and were thus excluded in the data analysis. Therefore, a total of 319 singleton mother–child pairs were included in the final analysis.

### Ethical Clearance

Ethical clearance was obtained from Kwame Nkrumah University of Science and Technology (CHRPE/AP/240/22). Informed written consent was obtained from the literate participants. In situations, where the participants could not write or read, thump-printed informed consent was obtained after providing the needed information and explanation. Additionally, respondents were provided with copies of the consented form.

## Results

### Socio-demographic characteristics of respondents

The total number of postpartum women enrolled was 322 but three of the completed questionnaires had some missing values and so 319 data set were included in the analysis. Table 1 shows the sociodemographic characteristics of study participants whose mean age was 29.4 ± 3.77 years, with more than half (61.8%) between the ages of 21 – 30 years. The majority (34.2%) were traders. In terms of education, 35.4% of them attained tertiary-level, 20.1% had JHS/Middle level education whilst 6% had no formal education. Almost all the respondents (99.4%) had only one child under two years and most of them (94.0%) were married.

**Table 1:** Socio-demographic characteristics of respondents (N = 319)

| **Variables** | **Frequency (n)** | **Percentage (%)** |
| --- | --- | --- |
| **Age of mothers (Years)** |  |  |
| Under 21 | 2 | 0.6 |
| 21 - 30 | 197 | 61.8 |
| More than 30 | 120 | 37.6 |
| **Marital status** |  |  |
| Married | 300 | 94.0 |
| Single | 19 | 6.0 |
| **Ethnicity of respondent** |  |  |
| Akan | 14 | 4.4 |
| Dagomba | 199 | 62.4 |
| Gonja | 57 | 17.9 |
| Others | 49 | 15.4 |
| **Educational Level** |  |  |
| Non formal education | 19 | 6.0 |
| Primary | 47 | 14.7 |
| JSS/Middle | 64 | 20.1 |
| SHS/Vocational | 76 | 23.8 |
| Tertiary | 113 | 35.4 |
| **Occupation** |  |  |
| Agriculture worker (e.g. farming) | 13 | 4.1 |
| Education/Research(teaching) | 51 | 16.0 |
| Health care (e.g. nursing) | 17 | 5.3 |
| Office worker (Civil servant) | 33 | 10.3 |
| Service worker (e.g. hair dresser) | 67 | 21.0 |
| Trader/Vendor | 109 | 34.2 |
| Unemployed | 19 | 6.0 |
| Others | 10 | 3.1 |
| **How many children under two years do you have?** |  |  |
| Only one child | 317 | 99.4 |
| Two children | 2 | 0.6 |

### 4.3 Medical and obstetric history of mother

Table 2 presents the obstetric and medical records of the respondents. Less than half (32.9%) of the respondents had malaria infection during pregnancy with index child. With regards to gravidity, 26.3 % were primigravida and just a little over half of the respondents (55.2 %) delivered through normal vaginal. Late/post (at least 41 weeks) delivery was by 23.5 % of respondents. The mean gestational weight gain (GWG) was low (151.7 g/week), with nearly all of the women (97.2 %) gaining less than the recommended American Institute of Medicine (IOM) GWG criteria.

**Table 2:** Medical and obstetric history of study participants.

| Variables | Frequency (n) | Percentage (%) |
| --- | --- | --- |
| **Frequency of malaria infection during pregnancy** |  |  |
| Zero | 214 | 67.1 |
| Once | 65 | 20.4 |
| Twice | 38 | 11.9 |
| Thrice | 2 | 0.6 |
| **Duration of pregnancy** |  |  |
| Term (37-40 weeks) | 211 | 61.1 |
| Pre-term (Less than 37 weeks) | 33 | 10.3 |
| Late/post (at least 41 weeks) | 75 | 23.5 |
| **How many months old was your pregnancy when you first attended ANC service?** |  |  |
| Three or less | 144 | 45.1 |
| Four above | 175 | 54.9 |
| **ANC attendance** |  |  |
| ≤ 8 | 50 | 15.7 |
| ≥ 8 | 269 | 84.3 |
| **How did you deliver?** |  |  |
| Caesarean section | 143 | 44.8 |
| Normal vaginal delivery | 176 | 55.2 |
| **Gravidity** |  |  |
| Primigravida | 84 | 26.3 |
| Secondi gravida | 71 | 22.3 |
| Multigravida | 164 | 51.4 |
| **First trimester BMI (kg/m<sup>2</sup>)** |  |  |
| Underweight (< 18.5) | 12 | 3.8 |
| Normal (18.5-25.0) | 125 | 39.2 |
| Overweight/obese (> 25.0) | 182 | 57.1 |
| **Mean maternal GWG per week (g)** |  |  |
| Inadequate, (<419.4) | 310 | 97.2 |
| Normal (419.4–692.3) | 9 | 2.8 |
| Excessive (>692.3) | 0 | 0.0 |

### Factors associated with gestational age at birth

The significant predictors of gestational age were weight in the first trimester, maternal height, marital status, age of mother, gestational rate of weight gain and malarial infection during pregnancy (Table 3).

**Table 3:** Predictors of gestational age at delivery (Multivariable regression analysis)

| Model | Standardized Coefficients | T | Sig. | 95.0% Confidence Interval for ($\beta$) | |
| --- | --- | --- | --- | --- | --- |
| | Beta ($\beta$) | | | Lower Bound | Upper Bound |
| (Constant) |  | 10.12 | <0.001 | 18.08 | 26.81 |
| Height of mother | 0.33 | 5.58 | <0.001 | 5.10 | 10.67 |
| Marital status of women (single) | 0.31 | 6.09 | <0.001 | 1.58 | 3.08 |
| Age of mother (years) | 0.28 | 4.09 | <.001 | 0.07 | 0.20 |
| Gestational rate of weight gain in grams per week | -0.12 | -2.38 | 0.018 | -0.005 | -0.01 |
| Malarial infection during pregnancy | -0.11 | -2.05 | 0.041 | -0.79 | -0.02 |
| Weight of mother in first trimester | -0.24 | -3.21 | 0.001 | -0.05 | -0.01 |

Older women were positively associated with longer gestational age as a unit increase in age of the mother increased gestational age by 0.28 standard units, β = 0.28, (95% CI: 0.07 to 0.20). Whereas maternal height positively associated with gestational age at delivery, β = 0.33, (95% CI: 5.10 to 10.67), weight of mother in first trimester associated negatively with gestational age at delivery, β = -0.24, (95% CI: -0.05 to -0.01). Malarial infection during pregnancy reduced duration of gestation by 0.11 standard units, β = -0.11, (95% CI: -0.79 to -0.02). Gestation of single women was longer than married women, β = 0.31, (95% CI: 1.58 to 3.08). Greater weekly gestational rate of weight gain reduced gestational age by 0.12 standard units, β = -0.12, (95% CI: -0.005 to -0.01)

The set of predictors accounted for 20.6 % of the variability in gestational age (Adjusted R Square = 0.206).

### Postnatal growth indicators according to gestational age at delivery

The results show that gestational age at delivery was associated positively with LAZ but negatively with weight for length (WLZ) (Table 4). Children born late-term/post term had significantly higher height-for-age z score than those born at term.

**Table 4:**
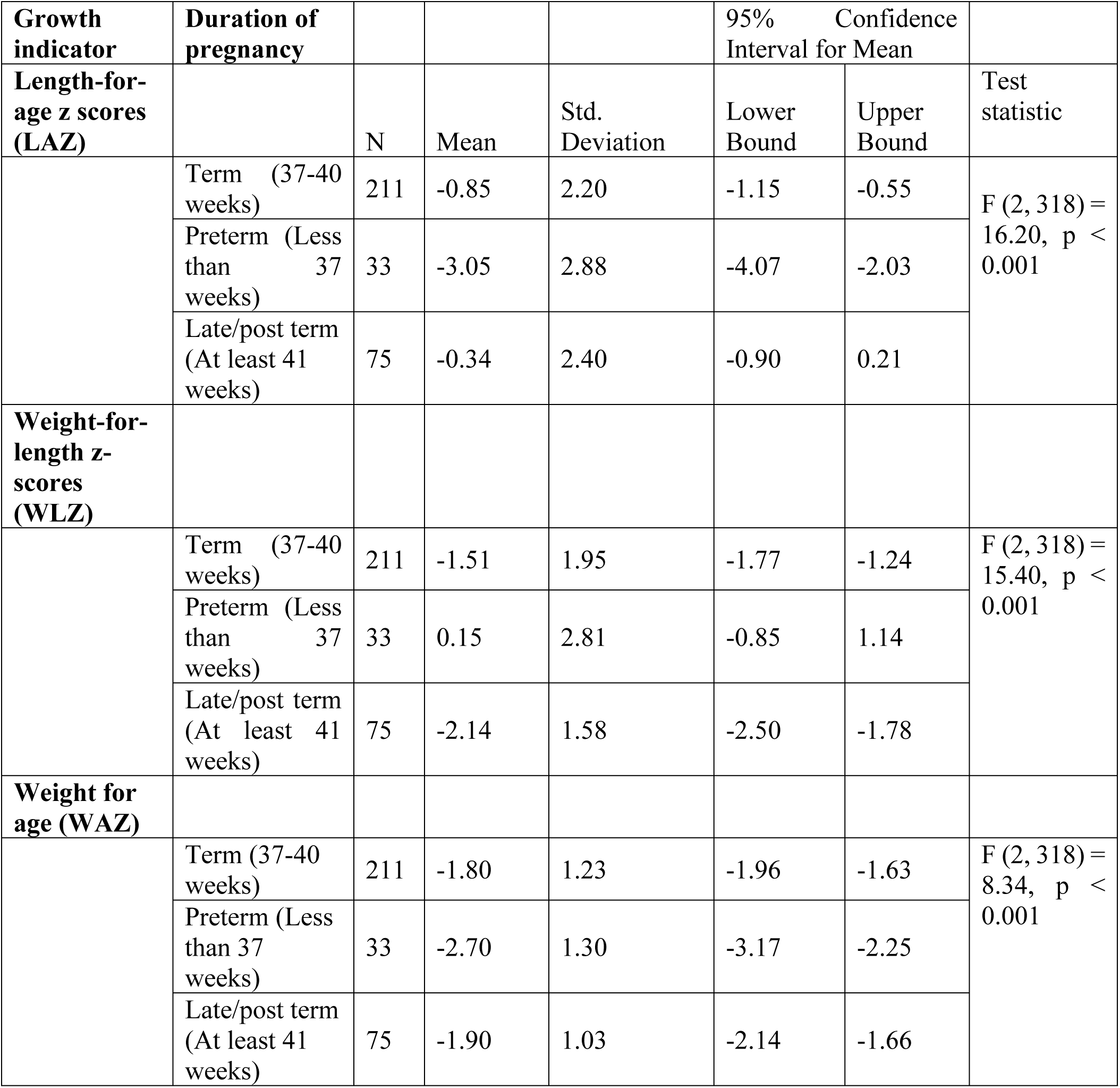
Child postnatal growth indicators according to gestational age at delivery (Bivariate analysis)

**Table 5:**
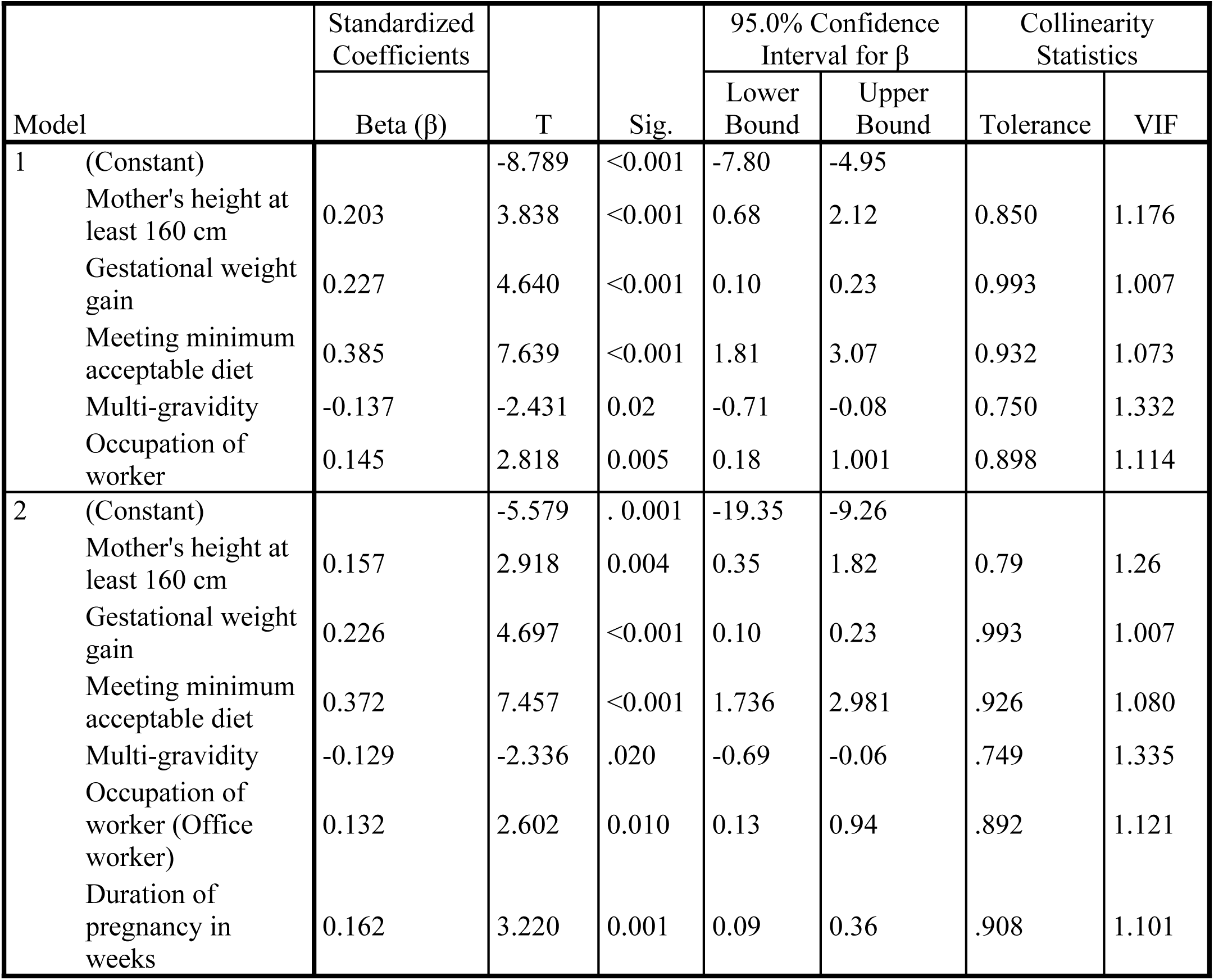
Predictors of LAZ (Multivariable regression analysis)

### Predictors of LAZ

In a hierarchical multivariable linear regression, we adjusted for confounders including birth weight, gender, age of child, maternal educational level, marital status, maternal age, ethnicity but these were significant.

An increase in one week of gestational age was associated with 0.16 increase in LAZ [beta coefficient (β), = 0.16 (95% CI: 0.09 to 0.36)]. Gestational weight gain had a significant positive effect on length-for-age z scores (LAZ), as a unit increase in the pregnancy weight was associated with an increase of 0.226 standard units in LAZ (β = 0.226, 95% CI: 0.10 –0.23). Children who were fed with minimum acceptable diet had a mean LAZ that was significantly higher by 0.372 standard units (beta β = 0.372, p < 0.001), compared to children who were not fed with it. Women who were office workers had higher LAZ, compared with their colleagues who were not workers ( β = 0.132, p = 0.001). Multi-gravida women children lower mean LAZ, compared with primigravida women (β = -0.129, p = 0.001).

Children of mothers whose height was at least 160 cm had higher LAZ, compared with their colleagues whose mothers height was less than 160 cm ( β = 0.157, p = 0.004). In step 2 of the regression analysis, when gestational age was added to the model, the % of variability accounted for went up from 24.6 % to 26.8 % (R^2^ Change = 2.4 %, p = 0.001). This means that, gestational age alone independently accounts for 2.4 % of the variance in LAZ.

### Predictors of stunting and wasting

A multiple logistic regression analysis showed variables that significantly predict stunting and wasting in preschool children aged 6–23 months (Table 6). Adjustment was made for potential confounding factors which included maternal educational level, marital status, maternal age, ethnicity, infant sex, birth weight, parity. However, these were not significant and were therefore removed from the model.

**Table 6:** Logistic regressions predicting odds of stunting and wasting.

| Variable | Stunting |  | Wasting |  |
| --- | --- | --- | --- | --- |
|  | COR (95% CI) | AOR (95% CI) | COR (95% CI) | AOR (95% CI) |
| **Duration of pregnancy in weeks** |  |  |  |  |
| Term (37-40 weeks) | Reference | Reference | Reference | Reference |
| Preterm (Less than 37 weeks) | 5.64 (2.59 – 12.26) *** | 2.93 (1.16-7.43)* |  | 0.92 (0.37-2.32) |
| Late/post term (At least 41 weeks) | 0.74 (0.38 – 1.43) | 0.80 (0.39 -1.64) |  | 2.18 (1.25-3.82)** |
| **Gestational weight gain** |  |  |  |  |
| At least 7 kg | Reference | Reference | Reference | Reference |
| Less than 7 Kg | 2.04 (1.06-3.95)* | 4.62 (1.94 -11.0) ** | 1.0 (0.60-1.68) | 1.0 (0.56-1.81) |
| **Met minimum acceptable diet (MAD)?** |  |  |  |  |
| Yes | Reference | Reference | Reference | Reference |
| No | 7.45 (4.0-13.87)*** | 6.43 (3.07-13.47) *** | 0.34 (0.18-0.65)** | 0.32 (0.16-0.67)** |
| **Age of child (years)** |  |  |  |  |
| 12-23 months | Reference | Reference | Reference | Reference |
| 6-11 months | 4.67 (2.66-8.22)*** | 2.96 (1.50 -5.84) ** | 0.32 (0.20-0.50)*** | 0.25 (0.14-0.44)*** |
| Maternal weight in first trimester | 0.96 (0.94-0.98)*** | 0.97 (0.95 -0.99) * | 1.007 (0.99-1.02) | 0.97 (0.95-0.99) ** |
| **Child had diarrhoea in the past two weeks?** |  |  |  |  |
| No | Reference | Reference | Reference | Reference |
| Yes | 0.94 (0.55-1.61) | 1 (0.52-1.93) | 1.68 (1.04-2.72) * | 1.73 (1.05-2.87) * |

\*significant at p <0.05; \*\*significant at p <0.01; \*\*\*significant at p <0.001
AOR (95% CI): Adjusted odds ratio at 95% confidence level
C OR (95% CI): Crude/unadjusted odds ratio at 95% confidence level

Stunting was 2.9 times more common among the children born through preterm AOR= 2.93 (95% CI 1.16, 7.43, and p = 0.03), compared to children born at term (37-40 weeks). Late/post term (At least 41 weeks) tended to protect against stunting in children AOR= 0.80 (95% CI: 0.39, 1.64, though this was statistically significant. Children who were not fed with minimum acceptable diet were 6.4 times more likely to be stunted than those who were fed with MAD (AOR: 6.43; 95% CI 3.07, 13.47, and p<0.001), Children whose mothers gained less than 7 kg during pregnancy were 4.6 times more likely to have stunted growth compared to children whose mothers gained at least 7 kg, AOR = 4.63 ( 95% CI: 1.94, 11.0, and p = 0.001). The set of predictors explained 35.6 % of the variance in stunting (Nagelkerke R Square = 0.356).

Wasting was 2 times more common among the children born late/post term, AOR= 2.18 (95% CI: 1.25, 3.82, and p = 0.006), compared to children born at term (37-40 weeks). Diarrhoea had no significant effect on stunting but children who had diarrhoea in the past two weeks prior to the study were 1.7 times more likely to be wasted, compared with their colleagues who had no diarrhoea, AOR= 1.73 (95% CI: 1.05, 2.87, and p = 0.03). Compared to older children (12-23 months), younger children (6-11 months) were 75 % protected against wasting AOR = 0.25, (95% CI: 0.14, 0.44, and p < 0.001). A unit increase in maternal weight in the first trimester provided 3 % protection against wasting AOR = 0.97 (95% CI:0.95-0.99, p = 0.009)

## Discussion

### Factors associated with gestational age at birth

The significant predictors of gestational age identified in this study were weight of mother in the first trimester, maternal height, marital status, older maternal age, gestational rate of weight gain and malarial infection during pregnancy. The study thus contributes to providing information that enriches an existing understanding of factors affecting the duration of gestation for women.

Older women were positively associated with longer gestational age as a unit increase in age of the mother increased gestational age by 0.28 standard units, β = 0.28, (95% CI: 0.07 to 0.20).

This finding is consistent with some other studies that have shown a positive association between older maternal age and longer gestational periods [21–23] but these findings are not consistent across all research.

One study found that increasing maternal age was associated with decreased spontaneous activity and an increased likelihood of polyphasic spontaneous myometrial contractions, which can prolong gestational length [24].

In this study, whereas maternal height positively associated with gestational age at delivery, weight of mother in first trimester associated negatively with gestational age at delivery. This means while taller mothers tend to have longer pregnancies (positive association between maternal height and gestational age at delivery), a mother’s weight in the first trimester is negatively associated with gestational age at delivery.

Our present findings indeed go to confirm earlier studies that suggest the risk of post-term deliveries increased with maternal height, meaning taller women tend to have longer pregnancies [23, 25, 26]. Late/post-term pregnancies were considerably less common in women who were shorter than 160 cm in height. They were 97.2 % less likely to have late/post-term pregnancy, as compared with women of tall stature. This implies that being shorter in stature helps prevent late or post-term babies. This could be ascribed to the fact that shorter women may have smaller pelvic dimensions, which could potentially lead to limitations in fatal growth or positioning. This could affect the baby’s ability to reach full term comfortably within the womb, reducing the likelihood of late-term or post-term pregnancies. Additionally, the size of the uterus may be correlated with maternal height. A smaller uterus might provide less space for the fetus to grow comfortably to full term, potentially leading to earlier labor.

Women who were underweight (low weight) in the first trimester are indeed more likely to have low gestational weight gain during pregnancy, and this can increase the age of delivery. This partly explains the negative association between a mother’s weight in the first trimester and gestational age at delivery.

Malarial infection during pregnancy reduced duration of gestation. Malaria infection during pregnancy can lead to miscarriage, premature delivery, low birth weight, congenital infection, and/or perinatal death. The exact mechanisms by which malaria causes these adverse outcomes are not fully understood, but they likely involve inflammation, placental dysfunction, and reduced oxygen and nutrient delivery to the fetus.

Gestation of single women was longer than married women. Greater weekly gestational rate of weight gain reduced gestational age. While the idea of increased weight gain leading to a shorter pregnancy might seem counterintuitive, it’s generally not the case because foetal growth is a process that continues throughout pregnancy, with significant gains occurring in the later stages, especially the third trimester. While weight gain can influence foetal size, it may not directly shorten the gestational period.

Gestational age is a non-modifiable factor but what can be done about long gestation? While gestational age is a non-modifiable factor, when a pregnancy extends beyond 40 weeks, healthcare providers offer options like elective induction of labor, expectant management with antenatal testing, or a combination of these strategies, depending on the individual circumstances

### Effect of late/post term pregnancy on stunting and wasting

In this analytical cross-sectional study, after adjusting for potential confounders, long duration of pregnancy associated positively with linear child growth as measured by LAZ. Post-term pregnancy was associated with a higher risk of thinness, but also protected against stunting. In a large population-based birth cohort study in China, post-term pregnancy was associated with a lower risk of overweight/obesity and a higher risk of thinness in preschool-age children [27]. Extended pregnancies can cause additional stress and anxiety for mothers, potentially impacting their ability to provide optimal postnatal care, including feeding practices, which can contribute to the infant’s risk of wasting.

The exact mechanisms behind these associations are not fully understood, but potential factors include the effects of prolonged gestation on foetal growth and development, as well as the timing of nutrient intake during pregnancy. These findings suggest that post-term pregnancies may have complex and potentially conflicting effects on the long-term growth and development of children aged 6–24 months.

The positive association between LAZ and gestational age at birth observed in this present study, has earlier been reported in studies conducted in Ethiopia Ghana, Malawi and Burkina Faso that found gestational age at birth to be strongly associated with child linear growth [28–30]. Though increased gestational age associated with LAZ, late or post term pregnancy is reported to be associated with other adverse perinatal and maternal outcome [13, 14] outcomes.

Aside long gestational period. the other main predictors of LAZ in children aged 6–24 months identified in the present study were gestational weight gain, child age, minimum acceptable diet, maternal height, and multi-gravidity. Identifying potentially modifiable factors associated with growth among children under two years of life remains a priority for the timely interventions. One of such factors identified in the present study is adequate gestational weight gain. Health care providers should therefore counsel pregnant women accordingly.

Maternal height was also a key predictor of LAZ where 1 cm increase in maternal height increased LAZ significantly. This finding is consistent with similar past studies conducted in low- and middle-income countries including Bangladesh, Brazil, India, Nepal, Peru, South Africa, and Tanzania s which found maternal height to have a significant impact on linear growth deficit up to 2 years of age [29–33].

## Conclusion

The prevalence of Late/post term delivery from the study was high at 23.5 % and it associated positively with linear child growth (protected against stunting) but was a high-risk factor for child thinness. These findings suggest that post-term pregnancies may have complex and potentially conflicting effects on the long-term growth and development of children.

## Strengths and limitations of the study

This study is significant because it’s one of the few in lower-income countries and the first in Ghana to examine how gestational age at birth impacts postnatal growth outcomes, addressing a gap in research in this area. The study fills a critical gap in research by focusing on the relationship between gestational age at birth and postnatal growth in lower-income countries, particularly in Ghana, where such research is scarce.

Despite its strengths, this study has several limitations. First, it was cross-sectional and therefore causality cannot be inferred. It does not capture potential changes in these factors over time. Longitudinal studies would be required to explore temporal trends and causality.

## Data Availability

All relevant data are within the manuscript and its Supporting Information files.

N/A

## Authors’ contributions

MS and CK conceived the study, participated in its design and contributed significantly to the acquisition of data. MS did the analysis and interpretation of data and together with CK drafted the manuscript and revised it critically for important intellectual content. Both the authors read and approved the final draft.

## Acknowledgements

The authors wish to acknowledge with gratitude the contribution of the data collection team whose hard work and commitment led to a successful conduct of this study. The co-operation and support of mothers and caregivers who took time off from their busy schedules to respond to the interviewers is very much appreciated.

## Notes

### Competing Interest Statement

The authors have declared no competing interest.

### Clinical Trial

N/A

### Clinical Protocols

N/A

### Funding Statement

The author(s) received no specific funding for this work.

### Author Declarations

Ethical clearance was obtained from Kwame Nkrumah University of Science and Technology (CHRPE/AP/240/22).

